# Automated Detection of Maternal Vascular Malperfusion Lesions of the Placenta using Machine Learning

**DOI:** 10.1101/2023.06.26.23291920

**Authors:** Purvasha Patnaik, Afsoon Khodee, Goutham Vasam, Anika Mukherjee, Sina Salsabili, Eranga Ukwatta, David Grynspan, Adrian D. C. Chan, Shannon Bainbridge

## Abstract

**Introduction:** Hypertensive disorders of pregnancy (HDP) and fetal growth restriction (FGR) are common obstetrical complications, often with pathological features of maternal vascular malperfusion (MVM) in the placenta. Currently, clinical placental pathology methods involve a manual visual examination of histology sections, a practice that can be resource-intensive and demonstrate moderate-to-poor inter-pathologist agreement on diagnostic outcomes, dependant on the degree of pathologist sub-specialty training.

**Methods:** This study aims to apply machine learning (ML) feature extraction methods to classify digital images of placental histopathology specimens, collected from cases of HDP [gestational hypertension (GH), preeclampsia (PE), PE + FGR], normotensive FGR, and healthy pregnancies, according to the presence or absence of MVM lesions. 159 digital images were captured from histological placental specimens, manually scored for MVM lesions (MVM- or MVM+) and used to develop a support vector machine (SVM) classifier model, using features extracted from pretrained ResNet18. The model was trained with and without data augmentation, and with and without data shuffling, and the performance of the classifiers assessed and compared through measurements of accuracy, precision, and recall using confusion matrices.

**Results:** The SVM model demonstrated accuracies between 7-78% for WSI-level MVM classification, with poorest performance observed on images with borderline MVM presence, as determined through post hoc observation.

**Conclusion:** The results are promising for the integration of ML methods into the placental histopathological examination process. Using this study as a proof-of-concept will lead our group and others to carry ML models further in placental histopathology.

## Introduction

The development and function of the placenta are essential to a healthy pregnancy outcome. When placental health becomes compromised, an umbrella of ‘placenta-mediated diseases’ can arise, including the most common obstetrical complications: hypertensive disorders of pregnancy (HDP) and fetal growth restriction (FGR) [1][2][3]. While the exact pathophysiology of these obstetrical complications is not entirely clear, a common finding of compromised utero-placental blood flow has been widely described in these conditions[1] [2][3]. Uterine spiral artery remodeling is vital for healthy pregnancy, and when disrupted, results in hypoxia-related placental injuries – observed visually as lesions of Maternal Vascular Malperfusion (MVM). It is estimated that ∼ 60% of all cases of HDP and FGR demonstrate some evidence of placental MVM lesions at the time of delivery [1][4][5] indicative of insufficient utero-placental blood flow as a contributory factor to the disease pathogenesis.

Over the years, perinatal pathologists have developed a detailed set of criteria that best determine the presence of MVM [6][7]. Microscopic placental lesions consistent with an MVM diagnosis have distinct visual appearances (Supplemental Figure 1) and include: placental infarcts, decidual arteriopathy, distal villous hypoplasia, accelerated villous maturation, increased syncytial knots, and villous agglutination [7]. Identification of MVM lesions upon placenta pathology examination can contribute to the appropriate diagnosis of placenta-mediated disease and serve as important components of maternal and neonatal care. Unfortunately, this clinical practice has historically been plagued by a lack of standardization in employed terminology and diagnostic criteria, is time and resource-intensive, and ideally must be performed by a specialty-trained perinatal pathologist – all collectively limiting the utility of this clinical practice [7][8]. Despite landmark efforts undertaken to improve global standardization in this field, the degree of inter-pathologist variability in examination findings remains high, particularly in low resource settings where specialized training and expertise are lacking [7]. Considering these challenges, placental pathology is a key clinical domain that may benefit most from the implementation of computer-aided diagnosis (CAD) through the development of machine learning algorithms capable of identifying distinct visual patterns in placental histopathology specimens [9], [10].

Computer vision is the ability of a computer to interpret and categorize images according to pre-defined labels, such as clinical diagnosis [11]. Through machine learning (ML), biomedical advances in computer vision have been progressing rapidly by using biomedical image datasets for segmentation and classification [12]. Supervised ML is the process of feeding a naïve ML model a dataset of images that have been pre-labeled for training [13]. The trained model is then deployed to label unseen and unlabelled images. If the model generalizes well (e.g., avoids underfitting and overfitting), it can operate with high accuracy on these new images.

As a step in this direction, the current study aims to develop and test supervised ML algorithms that can identify and classify digital images of placental histopathology specimens according to the presence or absence of MVM in cases of HDP and/or FGR, and healthy samples. A support vector machine (SVM) was developed to predict MVM lesions in whole-slide images of placental histopathology specimens. The results of this study will contribute to a small but growing body of work being carried out in this field [14]–[16], demonstrating the potential of CAD to improve the accessibility to consistent and high-quality diagnostic outcomes of the placenta pathology exam while reducing the resource-intensive workload of this important clinical practice. This can ensure that mothers and infants are receiving the best possible healthcare afforded to them.

## Methods

### Digital Image Dataset Acquisition and Patient Characteristics

This study received research ethics board approval from the University of Ottawa Research Ethics Board (#H-08-18-1023) and Mount Sinai Hospital (#13-0212-E). Original paraffin-embedded placenta histopathology specimens of villous tissue samples were purchased from the Research Centre for Women’s and Infants’ Health (RCWIH) Biobank at Mount Sinai Hospital (Toronto, ON). A total of 159 specimens were collected from patients with HDP [n= 17 gestational hypertension (GH); n = 45 preeclampsia (PE); n= 31 PE + FGR], normotensive FGR (n=20), and healthy controls (n=46), with each paraffin block containing four chorionic villous biopsies from each quadrant of the placenta. GH was defined as the onset of hypertension (systolic pressure ≥ 140 mm Hg and/or diastolic pressure ≥ 90 mm Hg) after 20 weeks of gestation, with the PE definition also requiring evidence of maternal end-organ dysfunction [17], [18]. FGR was defined as a neonatal birthweight <10^th^ percentile for gestational age and sex, with evidence of placental dysfunction in histopathology specimens [19]. Sectioned tissue was mounted on glass slides and stained following a standard histopathology protocol with Hematoxylin and Eosin (H&E). Slides were scanned at 20X magnification (0.5 µm/pixel) with an Aperio Scanscope digital scanner and saved in ‘.svs’ format.

### MVM Labelling of Images

All collated digital images underwent detailed histopathology examination by a perinatal pathologist (D. Grynspan, CHEO) blinded to clinical information, aside from gestational age at delivery. A synoptic data collection tool was used, which groups microscopic placental lesions according to broad placental disease categories [8]. The first category included in this tool is MVM, which includes the following lesions: placental infarcts, distal villous hypoplasia, accelerated villous maturation pattern, syncytial knots, and villous agglutination (Supplemental Figure 1)[7]. The pathologist scored each image for the absence (score = 0), presence (score = 1), and severity (score = 2; for applicable lesions only) of each MVM lesion identified. A cumulative MVM score was generated for the whole-slide image (WSI) by summing the pathology scores of all MVM lesions included in the synoptic tool (maximum score = 8). As healthy placentas may demonstrate mild evidence of one or more of these lesions, which may not reflect true placental disease, placentas with a cumulative MVM score ≥ 3 were classified as being diagnosed with MVM (MVM+). Placentas with a cumulative score between 0-2 were classified as MVM-, irrespective of clinical outcome. This classification threshold was determined based on the clinical expertise and guidance of the perinatal pathologist.

### Supervised Machine Learning Model Development

In this study, we developed a model to classify a WSI as MVM+ or MVM-, using a pretrained deep learning network for feature extraction and a support vector machine (SVM) for classification. The SVM model is commonly used in binary classification schemes, is easy to use in a clinical setting, and requires modest computing power for execution. The model was trained with and without data augmentation, and with and without data shuffling, and the results were compared. All coding used for image pre-processing, data augmentation, feature extraction, and SVM model development and testing was scripted in Python 3 language.

### Image Pre-processing

As a first step, the non-tissue part of each WSI was removed by producing a binary mask of the tissue. Images were converted to the HSV color space and Otsu thresholding was applied on the saturation channel. The output binary mask underwent morphological operations, such as opening and closing to remove small objects and fill small holes. A binary mask was applied to all WSIs to eliminate non-tissue parts of each digital image (Supplemental Figure 2).

As the size of each WSI is large (∼ 32,000 × 25,000 pixels), feeding the WSI directly into the model creates a high computational and memory load. Instead, a patch-based approach was employed, dividing each WSI into non-overlapping patches (1,500 × 1,500 pixels), with each patch given the same label as the source WSI’s label. Patches were then resized to 300 × 300 pixels. The total number of patches extracted from a WSI ranged from 23 – 228 patches, dependent on the amount of tissue present in the WSI. All patches were normalized using the singular value decomposition (SVD) geodesic method [20] to remove the color variation in H&E staining.

### Feature Extraction

For each patch, we extracted 512 features, using the pretrained ResNet18 network [21], from the average pooling layer before the fully connected layer. ResNet18 is a convolutional neural network that was pretrained on ImageNet [22], a large hand-annotated visual database of millions of images unrelated to histology (e.g., categories include items such as hummingbird, flamingo, and coffee mug). Each feature vector has an index for its source WSI, to enable WSI-level classification. At the end, these features along with their labels and indices were saved in hdf5 format to be used as an input for the SVM model.

### Model Development and Testing

a) Patch-level classification:

In this study, an SVM binary classification method was used to classify each patch as either MVM+ or MVM-, using a radial basis function kernel, gamma value of 0.001, and regulation parameter of 100.

b) WSI-level classification:

We used the majority voting method, across all the patches from a source WSI, to classify the WSI as either MVM+ or MVM-. In the majority voting method, the WSI was classified as MVM+ if at least the half of patches were classified as MVM+; otherwise, the WSI was classified as MVM-.

### Data Splitting and Augmentation

We performed 5-fold cross validation at the WSI-level, randomly splitting the WSIs into 5 partitions. For each fold, 80% of the WSIs were used for the training set and the remaining 20% were used for the test set, rotating the test set across the folds so that each WSI is eventually included in the test set. The total number of patches extracted from these 159 WSIs was 16,212, and the number of patches in a partition varied from 2,980 to 3,845.

The model was tested under four testing combinations: 1) no data augmentation and no data shuffling (i.e., original dataset), 2) no data augmentation, with data shuffling, 3) data augmentation, with no data shuffling, and 4) data augmentation and data shuffling. Data augmentation is used to artificially increase the amount of training data available, which can improve model performance [23]. Simple data augmentation types, including 90° rotations and image flipping, were performed on the WSIs prior to patch extraction which increased the training dataset by a factor of four. Data shuffling is used to avoid any ordering effects during training. It helps to produce a more generalized model [24]. Random WSI-wise shuffling was employed on the dataset.

### Classification Performance Metrics

Classification results for each of the proposed testing combinations, based on the ground truth labeling scheme of MVM+ and MVM-as scored by the blinded perinatal pathologist, were determined and compared. Classification performance metrics were visualized using confusion matrices to provide the ratios of true positives (TP), true negatives (TN), false positives (FP), and false negatives (FN) (Figure 1). Accuracy, precision, recall (sensitivity), and F1-score were calculated using prediction results on the test images, and specificity was manually calculated for each model. In addition, area under the curve (AUC) values were calculated, demonstrating how well the model can distinguish between the binary classes (MVM- vs. MVM+).

**Figure 1.**
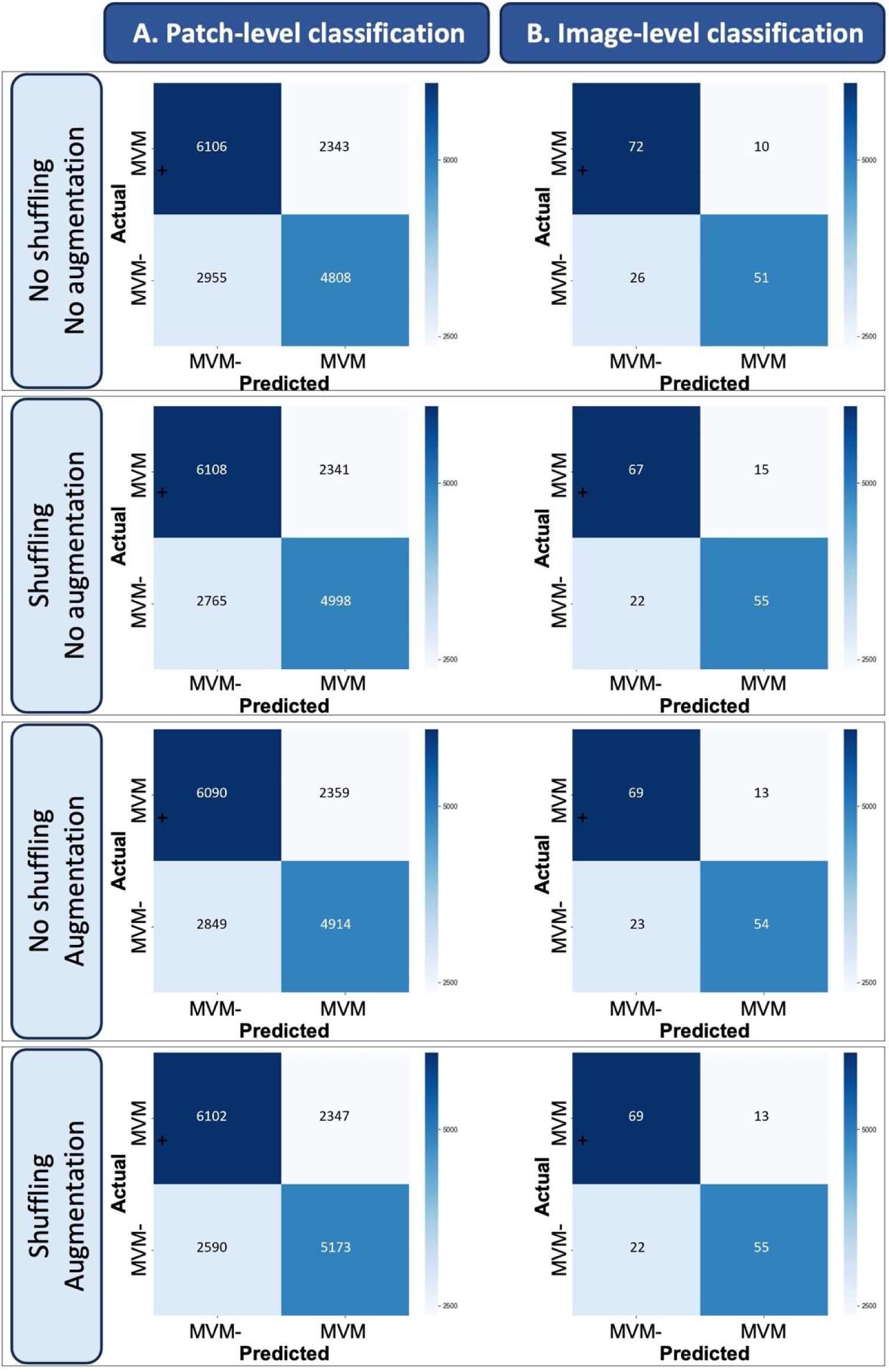
Confusion matrices for patch-level and WSI-level classification, with four different testing combinations, with or without shuffling and data augmentation. Quadrants in the confusion matrices represent true negatives (top left), false negatives (bottom left), true positives (bottom right), and false positives (top right). WSI = whole slide image, MVM = maternal vascular malperfusion lesion.

The classification performance metrics were calculated as follows:

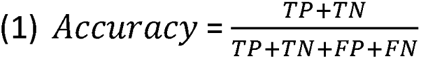

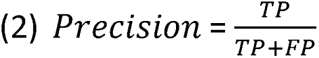

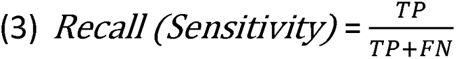

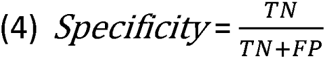

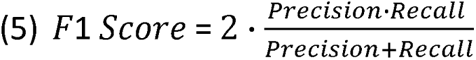

### Misclassification Analysis

A sub-analysis was performed to assess which sample characteristics were most associated with image misclassification, specifically examining the effect of pathologist derived MVM score (from 0 to 8), type of MVM lesion(s) present, and obstetrical diagnosis.

## Results

### Sample Demographics and Class Imbalance

Patient demographics according to pathologist derived MVM diagnosis are summarized in Table 1. There was a minor imbalance between the number of patients with MVM- (n=83) and MVM+ (n=76). Due to this imbalance and differences in the number of patches derived from each source WSI, a minor imbalance was also observed in the dataset regarding the total number of MVM- [n= 8449 (52%)] and MVM+ [n=7763 (48%)] patches (Supplemental Table 1).

**Table 1.**
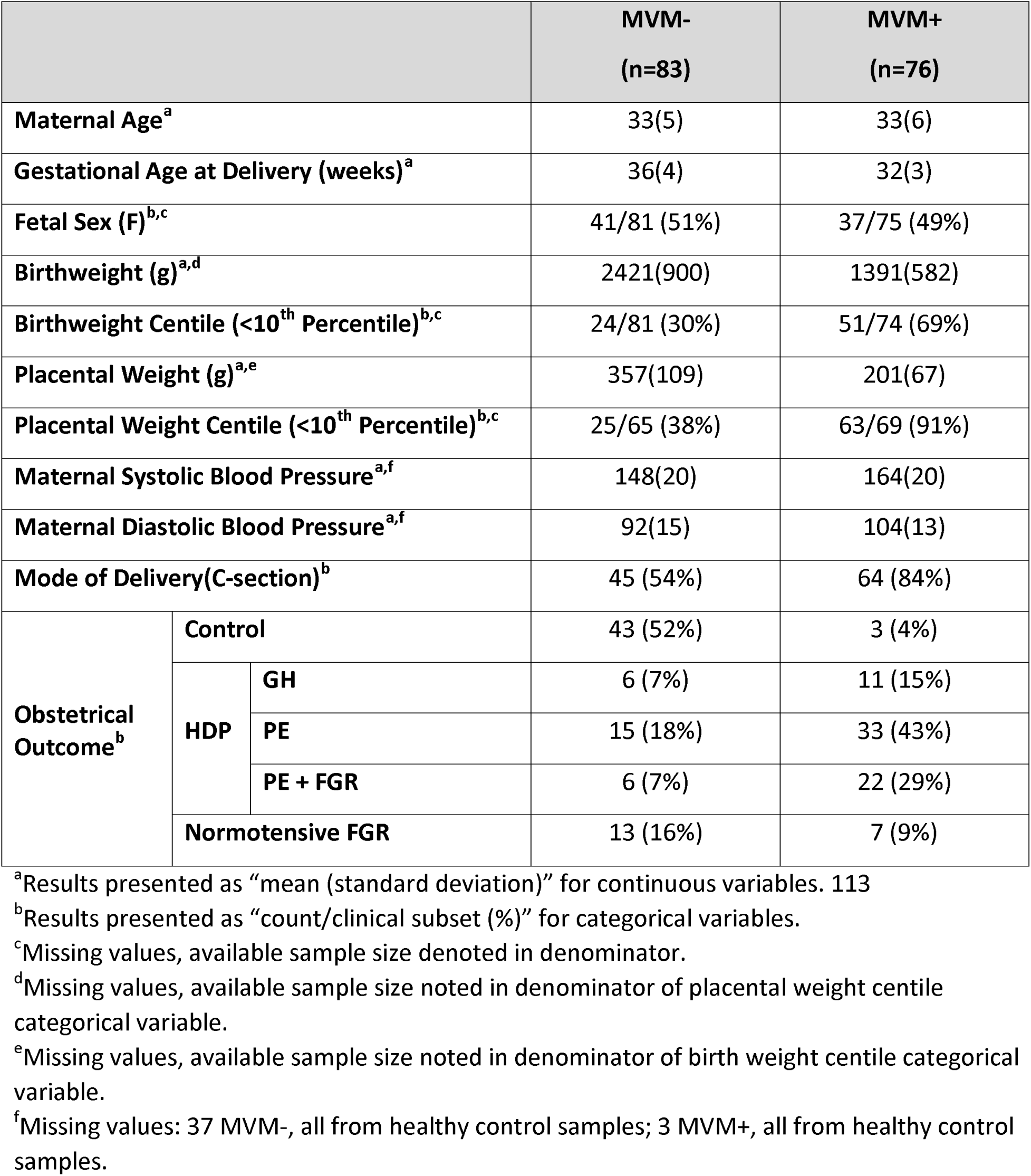
Demographic characteristics of participants in the study.

### Classification Performance Metrics

An SVM model which used a ResNet18 pretrained feature extraction method, produced accuracies ranging from 67-70% for patch-level and 77-78% for WSI-level MVM classification (Figure 1; Table 2). Results suggest that data augmentation and data shuffling can increase model performance, with the highest classification performance achieved when both are employed (accuracy = 78%, F1-score = 76%, AUC = 87% in WSI level; Figure 2; Table 2).

**Figure 2.**
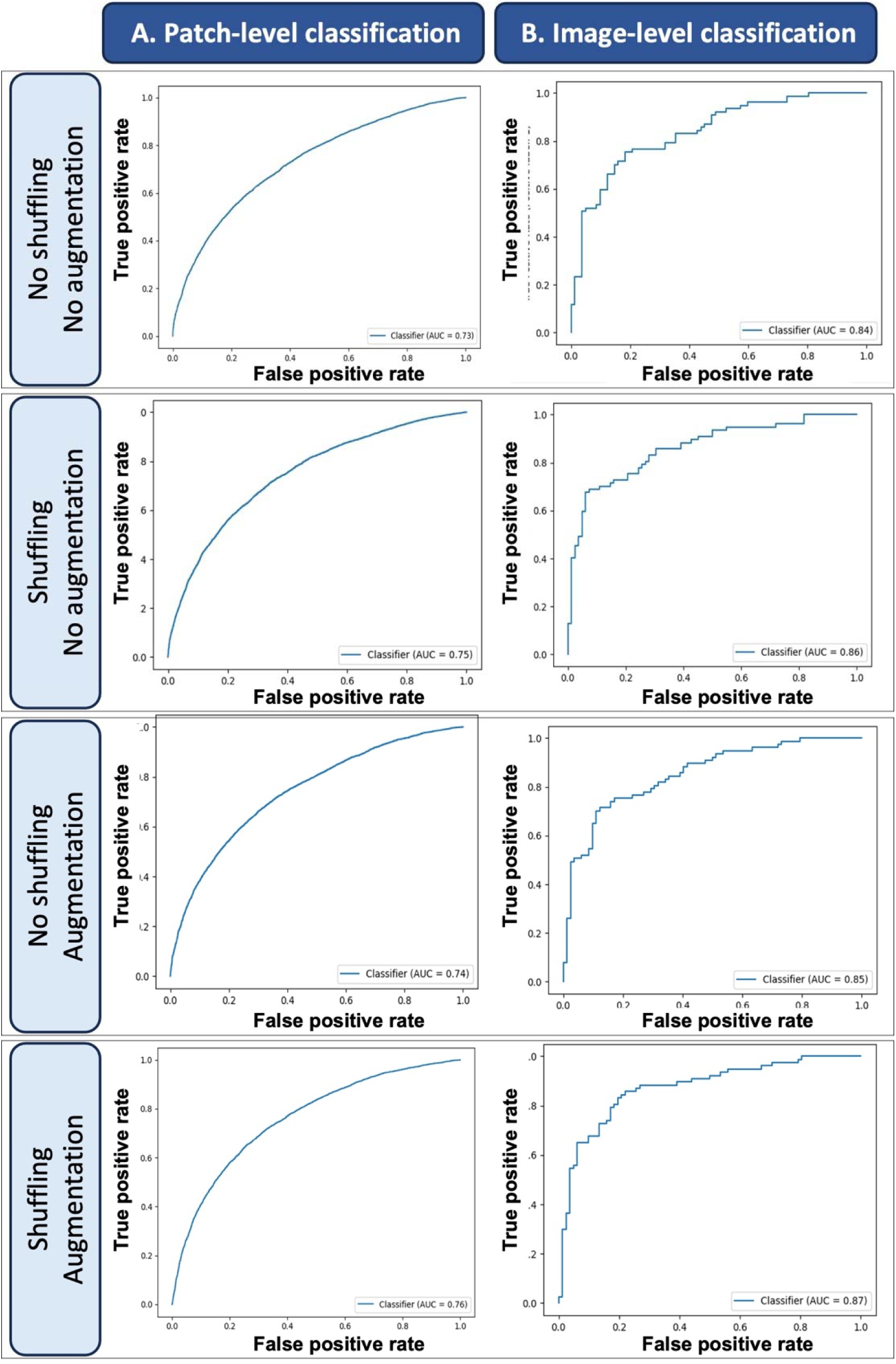
ROC curve for patch-level and WSI-level classification with four different testing combinations, with or without shuffling and data augmentation. ROC = Receiver operating curve, WSI = whole slide image.

**Table 2.** Summary of classification performance metrics for the developed model, with four combinations

| Testing combinations | Classification Model | Accuracy | Precision | Recall (Sensitivity) | Specificity | F1-Score | AUC of ROC |
| --- | --- | --- | --- | --- | --- | --- | --- |
| No shuffling + | Patch-level | 0.67 | 0.67 | 0.61 | 0.72 | 0.64 | 0.73 |
| No augmentation | WSI-level | 0.77 | 0.83 | 0.66 | 0.87 | 0.74 | 0.84 |
| Shuffling + | Patch-level | 0.68 | 0.68 | 0.64 | 0.72 | 0.66 | 0.75 |
| No augmentation | WSI-level | 0.77 | 0.77 | 0.71 | 0.82 | 0.75 | 0.86 |
| No shuffling + | Patch-level | 0.68 | 0.68 | 0.63 | 0.72 | 0.65 | 0.74 |
| Augmentation | WSI-level | 0.77 | 0.81 | 0.70 | 0.84 | 0.75 | 0.85 |
| Shuffling + | Patch-level | 0.70 | 0.69 | 0.67 | 0.72 | 0.68 | 0.76 |
| Augmentation | WSI-level | 0.78 | 0.81 | 0.71 | 0.84 | 0.76 | 0.87 |

### Image Misclassification

Across the MVM pathology score spectrum (0-8), model misclassification of images was similar for all testing combinations (Figure 3). A normal distribution in image misclassification was observed across MVM scores of 0-7, with the highest degree of image misclassification observed with MVM pathology score of 3 - the diagnostic threshold used for the MVM+ label designation. Cases with an MVM score of 8 (n = 3), representative of a high degree of placenta pathology, also demonstrated a moderate degree of misclassification with all 4 testing combinations. No difference in model misclassification was observed when a sub-analysis was performed according to the types of MVM lesions present (Supplemental Figure 3), however all cases of villous agglutination (n=3) were correctly identified (accuracy = 100%).

**Figure 3.**
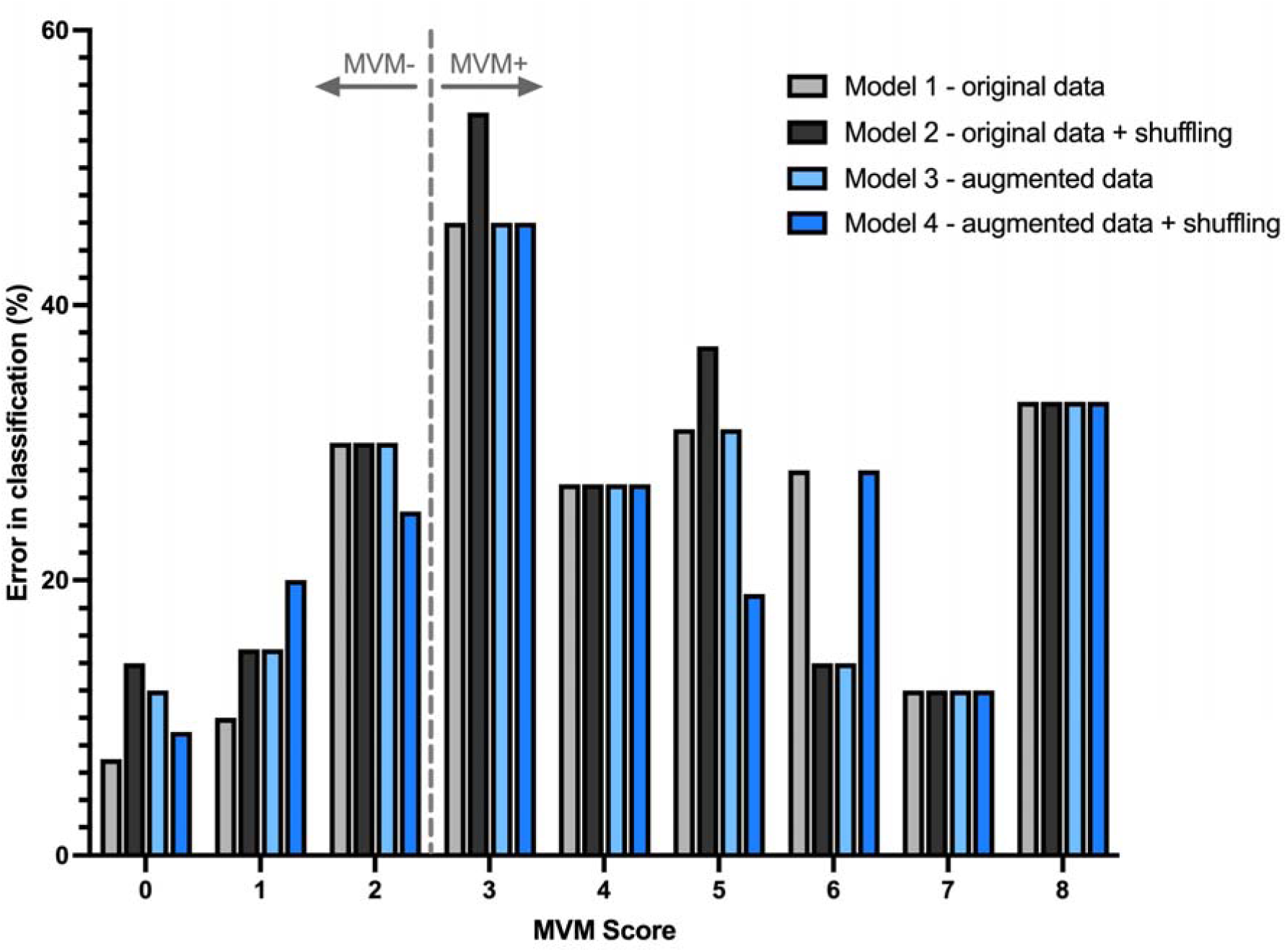
Misclassification error of images according to MVM pathology score. MVM score of ≥ 3 was used as the diagnostic threshold for the MVM+ label designation. MVM = maternal vascular malperfusion lesion.

WSI from healthy controls, with MVM pathology scores ranging 0-3 (Figure 4A), had the highest degree of misclassification at the MVM diagnostic threshold of 3 (Figure 4B-C). The false negative rate (missed cases) was highest for control patients, likely driven by control cases with an MVM score of 3 (Supplemental Figure 4). Unsurprisingly, HDP cases had higher overall MVM pathology scores compared to healthy controls (Figure 4A), but likewise demonstrated the highest number of misclassified images when the MVM pathology score hovered around the MVM+ diagnostic threshold (Figure 4B, D). When broken down according to the type of HDP, PE and PE + FGR cases specifically demonstrated this trend, whereas misclassification of GH was equally distributed across MVM pathology scores (Supplemental Figure 5). The false positive rate (false alarms) was highest for cases of HDP, likely driven by those cases that had low MVM scores but may have had other pathological forms of placental disease. This likely speaks to the high degree of heterogeneity of placental disease in cases of HDP that extend beyond MVM, and possibly the inclusion of cases with GH.

**Figure 4.**
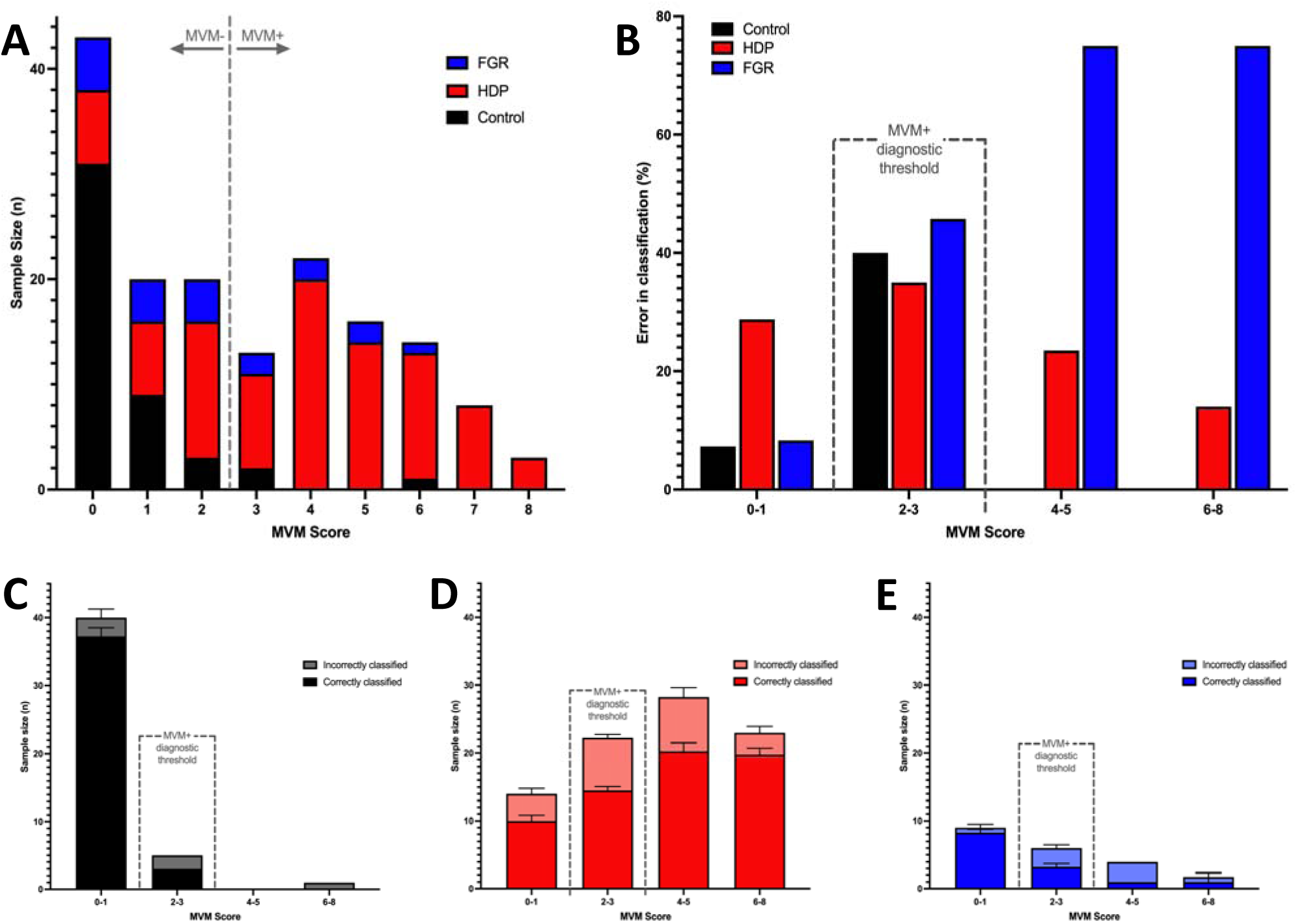
Model classification according to obstetrical outcome and MVM pathology score. **A**) Distribution of samples from each obstetrical outcome group, according to MVM score, **B**) Mean misclassification error across all models tested according to obstetrical outcome, **C**) Breakdown of accurately classified and misclassified control samples, according to their MVM pathology score, **D**) Breakdown of accurately classified and misclassified HDP samples, according to their MVM pathology score, **E**) Breakdown of accurately classified and misclassified FGR samples, according to their MVM pathology score. An MVM score of ≥ 3 was used as the diagnostic threshold for the MVM+ label designation. MVM = maternal vascular malperfusion lesion; HDP = hypertensive disorder of pregnancy; FGR = fetal growth restriction.

Interestingly, normotensive FGR cases had a wide range of MVM pathology scores (0-6; Figure 4A) and demonstrated the highest rates of WSI misclassification at higher MVM scores (4-8; Figure 4B, E), however the number of total cases with these scores was low (n=5; Figure 4A). The normotensive FGR cases demonstrated an equal rate of false negatives and false positives (Supplemental Figure 4).

## Discussion

In the current proof-of-concept study, an SVM model with 4 different testing combinations was evaluated for the classification of the presence/absence of MVM lesions in a WSI dataset of placental histopathology images. MVM lesions in the placenta are indicative of poor utero-placental perfusion across pregnancy and are common in cases of HDP and FGR. The developed model, which employed a pretrained ResNet18 feature extraction method, demonstrated promising classification performance which was further enhanced by the combined use of data augmentation and data shuffling.

Under the optimal testing combinations, that employed data shuffling and augmentation and performed classification at the WSI-level using majority voting, classification accuracy was 78%. Usurpingly, across the entire dataset, the misclassified WSIs often had a cumulative MVM score that hovered around the manual MVM+ diagnosis threshold established for this study. This ‘ground truth’ MVM+ diagnosis threshold used to convert a continuous MVM score variable into binary classes (cumulative score ≥3) was selected by a highly trained perinatal pathologist, based on his clinical expertise and experience. And while this threshold did often coincide with a relevant obstetrical outcome of HDP or FGR (in 96% of MVM+ cases, Table 1), placental disease demonstrates a spectrum of lesion presence and severity, with increasing pathology more likely associated with adverse impacts on the mother and fetus. As such, it is recognized that the selected threshold is subjective, and it is therefore not surprising that those WSIs with scores surrounding this threshold were more likely to be misclassified. Future work in this area should specifically address this study limitation, either through comparison of model performance at different MVM+ thresholds and/or moving past a binary classifier, rather attempting to develop a multi-class model that may better capture the spectrum of pathology seen clinically.

When assessing the performance of these models, one must also consider the issue of heterogeneity of pathology across the placenta. Some MVM lesions, such as distal villous hypoplasia, are often present in a diffuse pattern across the placenta, and hence will be present in both the original WSI and most image patches generated from the source WSI and used for model training. However, other MVM lesions, such as placental infarcts, may be more localized in nature. While the source WSI may be originally classified as MVM+ due to the presence of this lesion, microscopic evidence of this lesion may not be present in all, or even many, of the generated patches used to train the models. In fact, any lesion which demonstrates a high degree of heterogeneity across the placenta, and hence across the source WSI, will give rise to a larger number of mislabeling image patches, leading to poorer training and hence performance of the developed model. In the current study, WSI-level classification was achieved using a simple majority voting. This appeared to be sufficient for our dataset, as similar rates of WSI-level misclassification (∼20%) were observed when the placentas contained a diffuse/homogenous presenting MVM lesion such as DVH and more localized/heterogenous presenting MVM lesion such as placental infarct (Supplemental Fig 1). However, future research in this area should explore the use of attention-based, self-supervised learning approaches to mitigate any potential effects of lesion heterogeneity and patch mislabelling.

The results of the current study certainly highlight the potential utility of ML models to automate aspects of placental histopathology evaluations, while identifying aspects of image pre-processing and model optimization that can be explored further to bring this modality to the forefront. This study adds to a limited, but rapidly growing, body of work in this area [14]– [16] that collectively aims to develop, improving and validate ML models for placenta histopathology applications, with a concerted goal to optimize this CAD-modality for integrated clinical use.

### Limitations

The success of ML model development for use with clinical imaging data has in large part been driven by access to robust training datasets. In domains of clinical pathology, where the biggest gains in ML implementation have been made (i.e., onco-pathology), successful model development has been achieved using training datasets that include 500 – 10,000 original WSIs, which have then been augmented for training purposes [20], [25], [26]. Comparatively, the digital image sample sizes used in the current study are quite small (159 WSIs). Compounding the issue of sample size is the high degree of heterogeneity in clinical presentation and underlying placental pathology in cases of HDP and FGR. Poor utero-placental perfusion, as evidenced by the presence of MVM lesions, is considered a key component of disease pathophysiology for these conditions, particularly in cases of PE +/- FGR. However, it is important to note that not all cases of HDP or FGR demonstrate evidence of MVM (including ∼ 35% of the HDP and FGR cases included in the current study; Table 1), indicating that other forms of disease pathophysiology can lead to these obstetrical outcomes. For example, we have previously demonstrated the presence of distinct subclasses of PE and FGR, whose disease pathophysiology is instead driven by pro-inflammatory processes or maternal pre-disposing factors [18][19][27][28]. Importantly, multiple forms of placenta pathology can co-exist in these conditions, presenting clinically with the co-occurrence of MVM with others types of placenta lesions [i.e., villitus of unknown etiology (VUE), fetal vascular malperfusion (FVM) etc.]. The presence of these additional lesions, which also demonstrate unique visual features, may serve to distort MVM-labelled feature extraction and limit classification performance. The sub-analysis of image misclassification according to obstetrical outcomes performed in the current study certainly points to these limitations. This was particularly true for our GH and FGR cases, where image misclassifications tended to span the MVM score spectrum (Figure 4, Supplemental Figure 5), rather than being localized around the threshold value as seen with the other obstetrical outcome groups. To address the above-mentioned limitations, the clinical and research communities must work together to generate, and make available for research purposes, a robust clinical image dataset of reproductive tissues, including the vital organ of pregnancy - the placenta. Ideally, the inclusion of lesion-annotated datasets within such a research-ready image database would allow for timely and robust ML model development to address clinical needs unique to the health and wellbeing of mothers and their offspring.

### Future Work

The current study not only shows promise in using supervised ML in placental pathology but also uncovers important next steps. While continued work to further advance traditional ML models such as those tested in the current study is warranted, additional investigation into the use of deep learning models may also yield important contributions to this field. A move from transfer learning for feature extraction, as done in the current study, to using end-to-end convolutional neural networks (CNN) may prove to be an ideal model for histopathology image classification with high accuracy [29]–[31]. However, it is important to consider that each model has its pros and cons; where CNNs are prevalent in object recognition, they requires a much larger dataset than SVMs, whereas SVMs are highly prevalent in classification analysis [32]. Therefore, testing and comparing different paradigms in computer vision such as classical ML (e.g., SVM, random forest), deep learning (CNNs) with and without transfer learning (ex. VGG-16, ResNet-50) is essential to gaining perspective on the most optimal methods for placenta histopathology classification. The ability to automate the detection of MVM and other forms of placental disease would open the door to the integration of ML approaches into the placental pathology workflow in the future.

#### Author Contributions

Conceptualization, SAB, ADCC, DG; consultation on model development and coding, GV, SS, EU; data collection and analysis, PP, AK, GV, AM; writing, PP, AK, SAB; review and editing, EU, ADCC, SAB.; project supervision and funding, ADCC and SAB All authors have read and agreed to the published version of the manuscript.

#### Funding

This work was supported by the Canadian Institute of Health Research (CIHR; #CPG-170604, awarded to SAB and ADCC; #PJT-153055, awarded to SAB) and the Natural Sciences and Engineering Research Council of Canada (NSERC; #CHRP-549538-20, awarded to SAB and ADCC).

## Data Availability

All data produced in the present work are contained in the manuscript.

## Supplemental Figure Legends

**Supplemental Figure 1.**
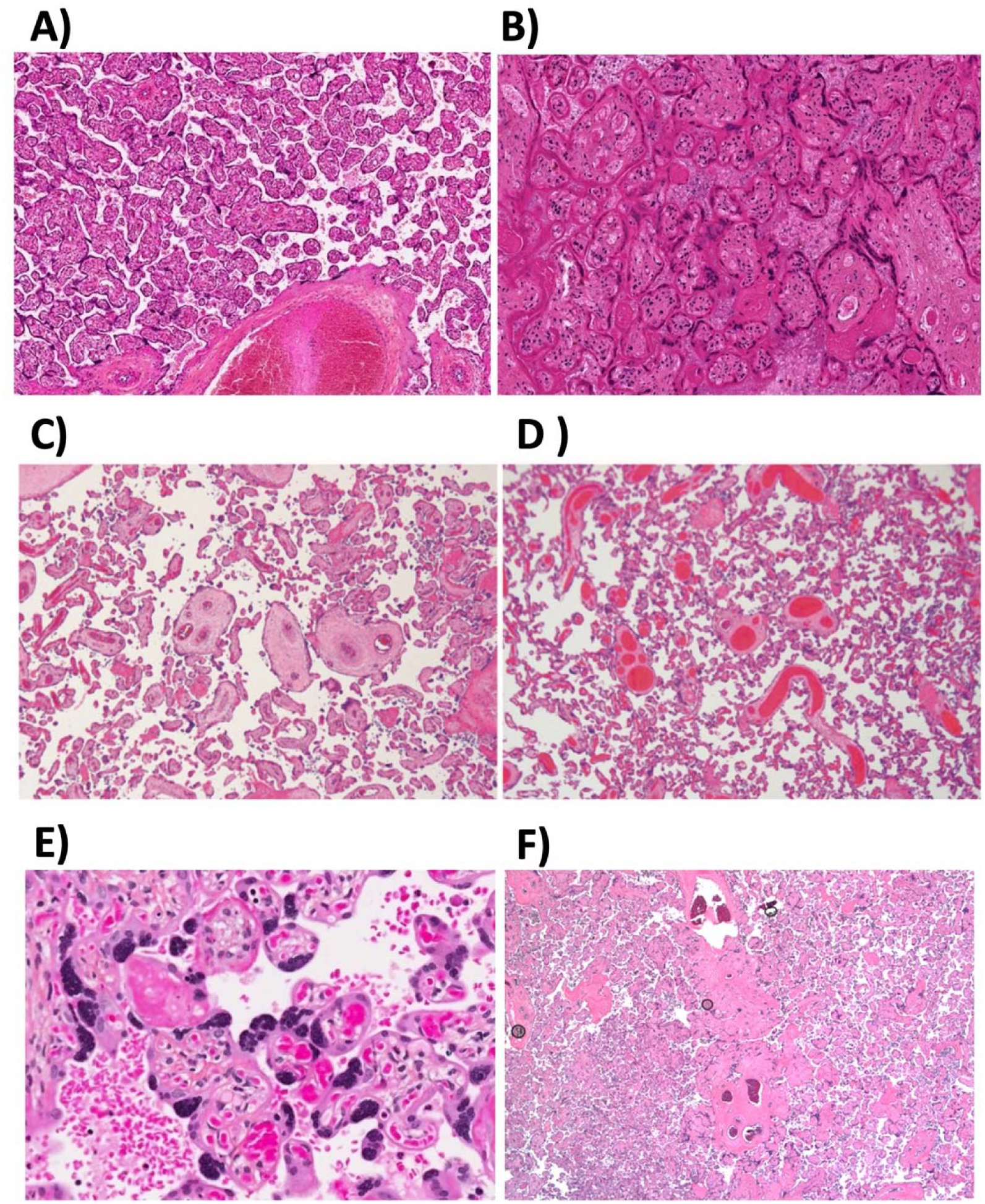
Placental lesions that define MVM. AA Normal placenta at term, B) Placental infarct, C) Distal Villous Hypoplasia, D) Accelerated Villous Maturation, E) Syncytial knots, F) Villous Agglutination. (Representative images provided by Dr. David Grynspan). MVM = maternal vascular malperfusion lesion.

**Supplemental Figure 2.**
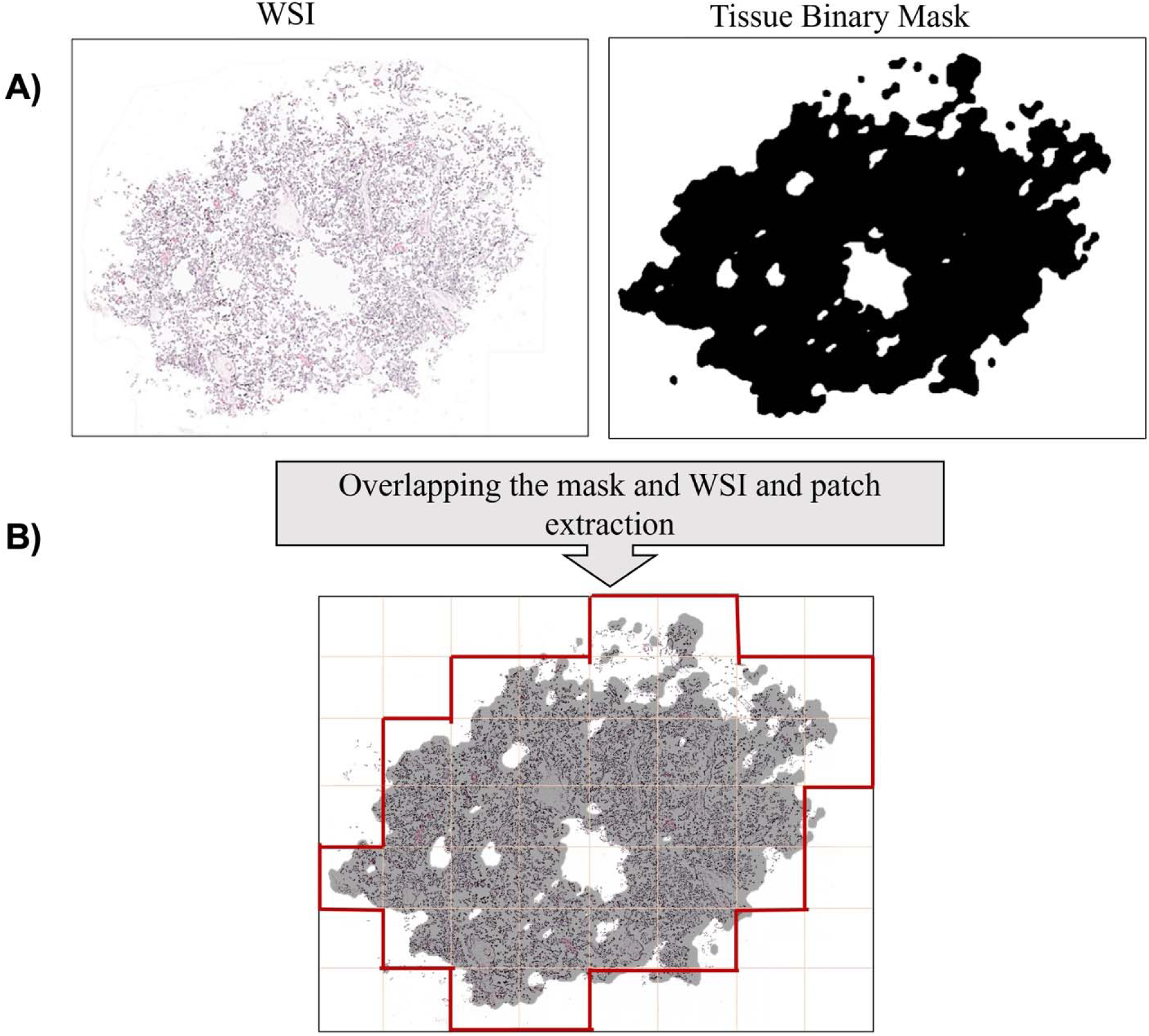
Tissue extraction and Patch Extraction. Creating binary masks to include the tissue section of slide to extract patches. WSI = whole slide image

**Supplemental Figure 3.**
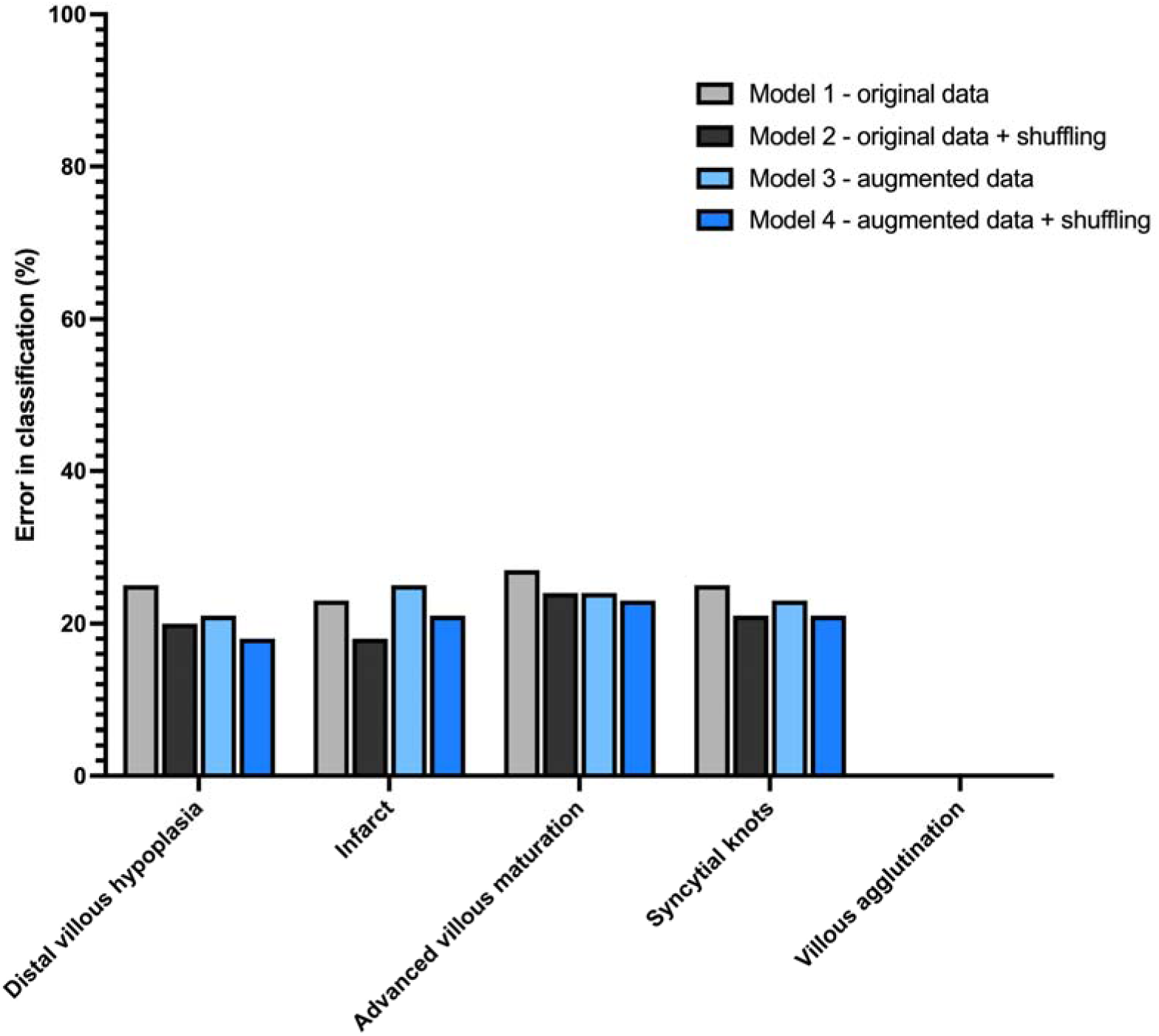
Misclassification error of images for each model tested according to the presence of distinct types of MVM lesions. MVM = maternal vascular malperfusion lesion.

**Supplemental Figure 4.**
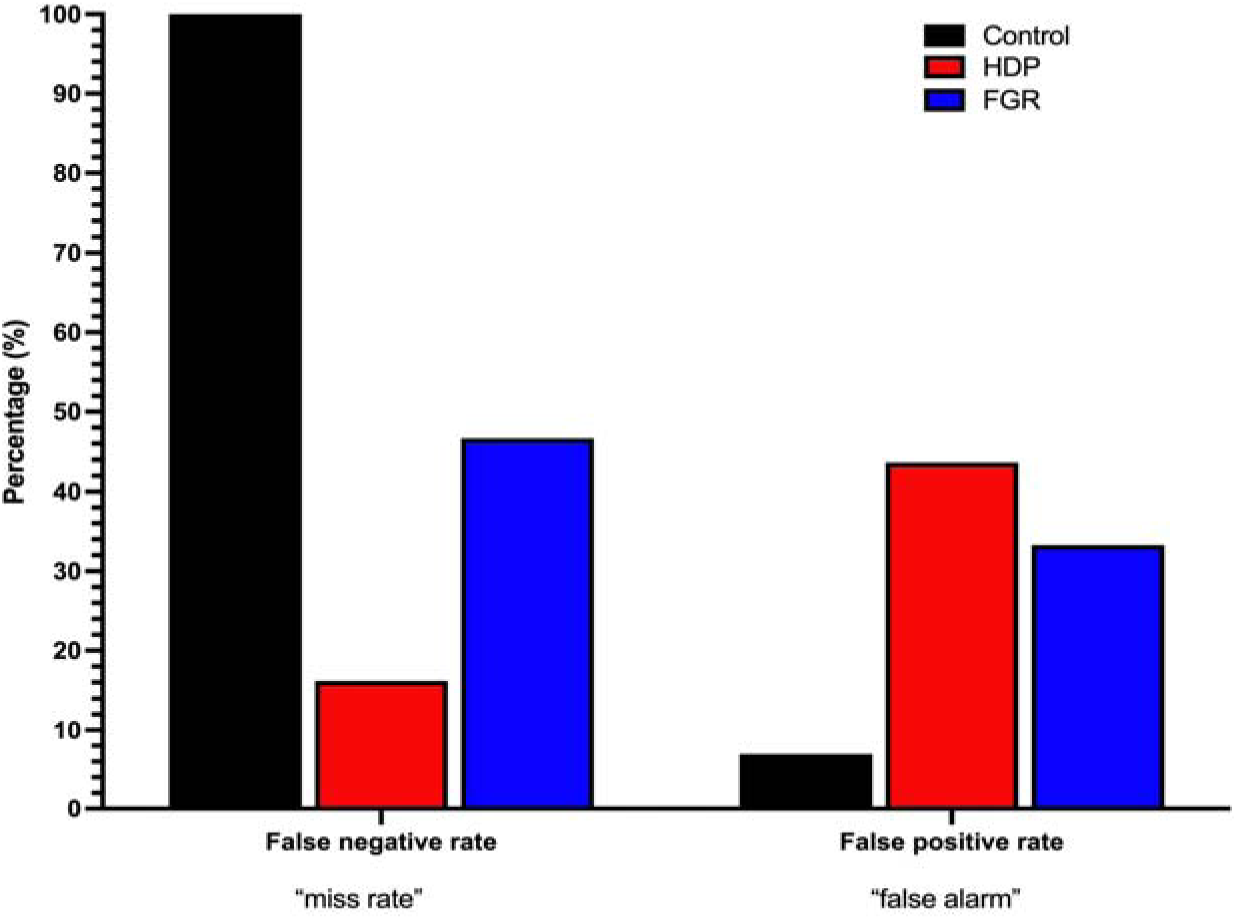
Mean false negative and false positive rates of models tested according to obstetrical outcome group. The false negative rate refers to how often the model incorrectly predicts the negative class– “missed rate”. The false positive rate refers to how often the model incorrectly predicts the positive class – “false alarm”. HDP = hypertensive disorder of pregnancy; FGR = fetal growth restriction.

**Supplemental Figure 5.**
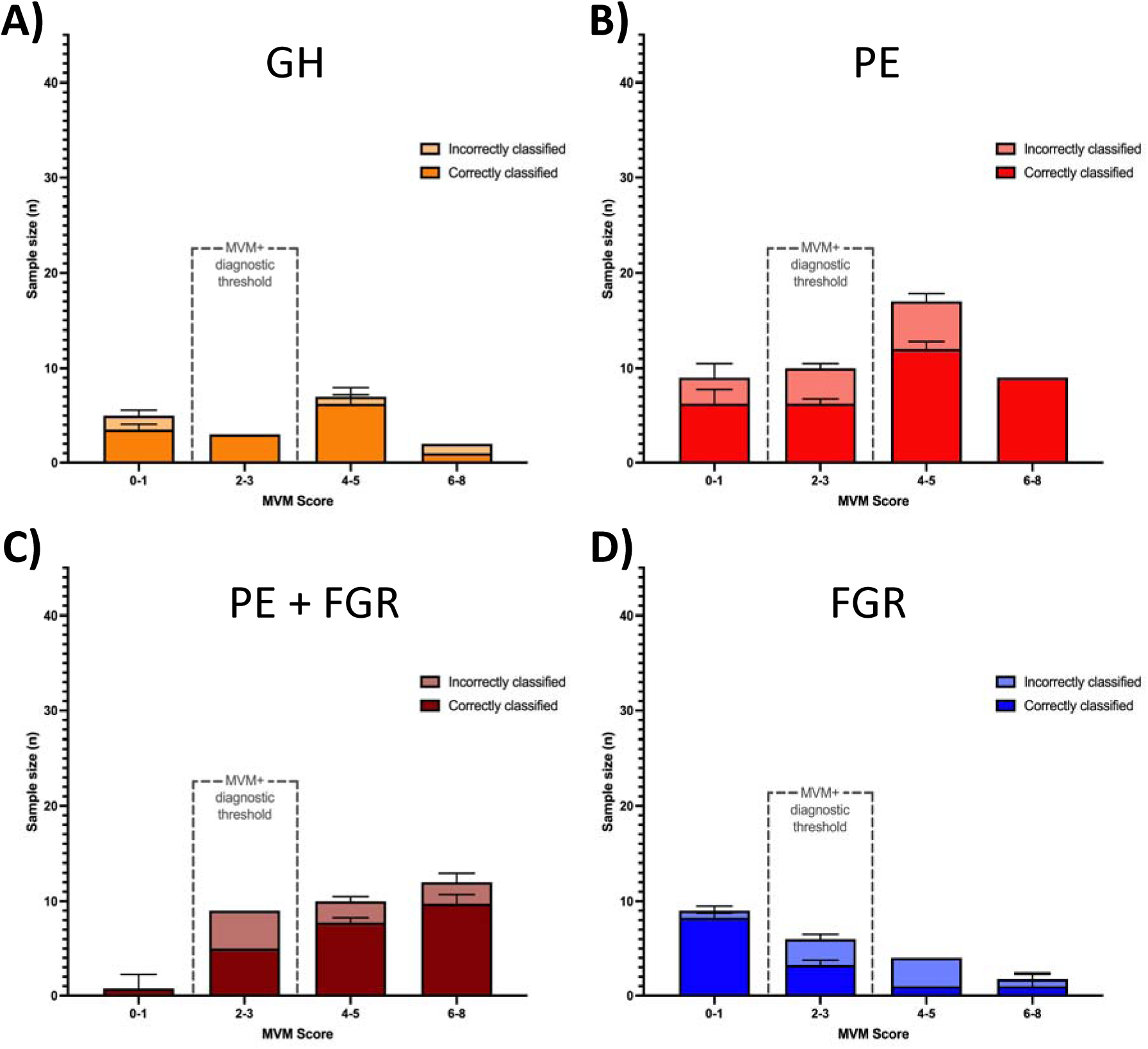
Breakdown of accurately classified and misclassified samples, according MVM pathology score and obstetrical outcome group, including: hypertensive disorders of pregnancy, broken down into PIH (A), PE (B) and PE + FGR (C); and, normotensive FGR (D). An MVM score of ≥ 3 was used as the diagnostic threshold for the MVM+ label designation. MVM = maternal vascular malperfusion lesion; PIH = pregnancy induced hypertension; PE = preeclampsia; FGR = fetal growth restriction.

**Supplemental Table 1.**
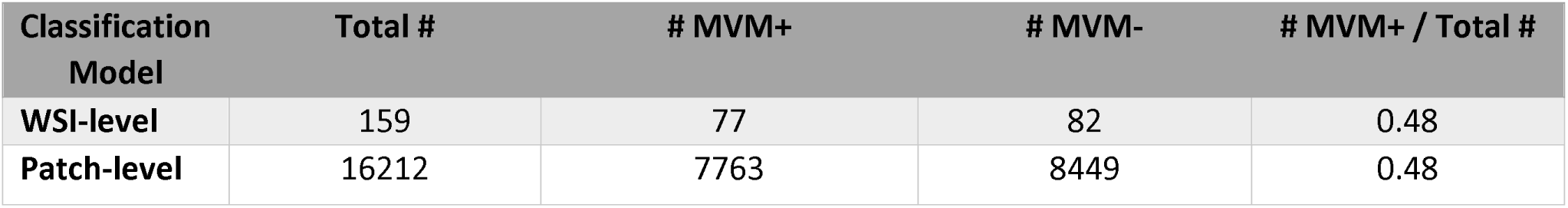
Summary of dataset size

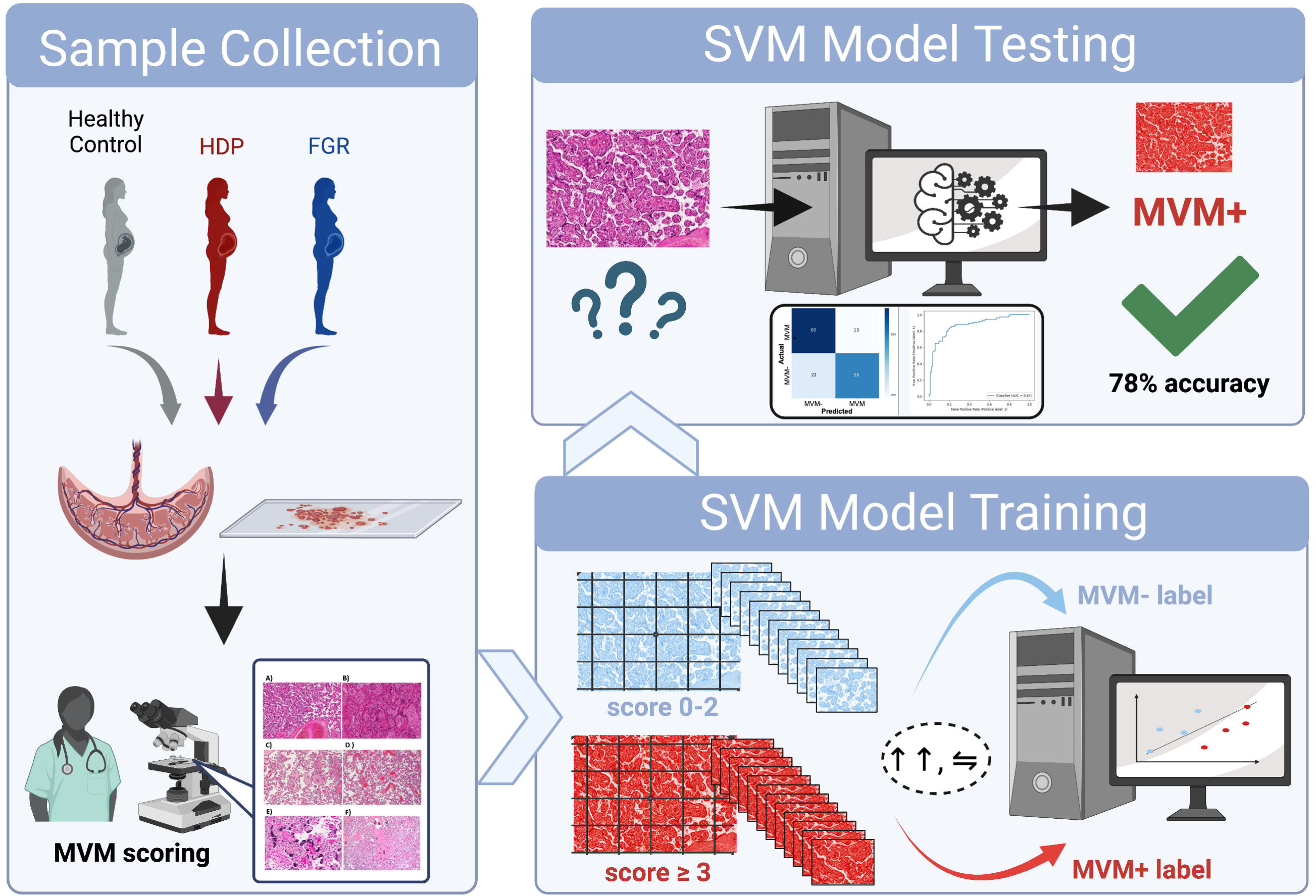

